# Attendances, Feeding Practices and Weight Trajectory of a Rural Cohort of Infants from Birth to 9 Months of age at a Facility-Based Well-Child Clinic

**DOI:** 10.1101/2024.10.30.24316420

**Authors:** Adenike Oluwakemi Ogah

**Affiliations:** Department of Pediatrics and Child Health, School of Medicine, University of Zambia, Lusaka, Zambia

**Keywords:** weight faltering, increment versus static/serial weight assessments, frequency of feeding, exclusive breastfeeding, clinic attendances, sex, cow milk, soya milk, full exclusive breastfeeding, supplementary and complementary feeds

## Abstract

**Background:** The identification of specific age windows and targeted population subgroups is essential for directing future preventive interventions aimed at addressing infantile weight faltering during the initial critical 1,000 days of life. These efforts are crucial as they have the potential to significantly impact both short-term and long-term health outcomes and overall survival.

**Subject and methods:** This study constituted a secondary cross-sectional analysis of data derived from a prospective cohort study. The data encompassed 529 mother-newborn pairs, who were enrolled at birth and subsequently monitored for a period of 9 months at the well-child clinic. The study evaluated infant attendances, feeding patterns and weight growth.

**Results:** At birth, there were 113 (21.4%) small-for-gestational-age, 379 (71.6%) appropriate-for-gestational-age and 37(7.0%) large-for-gestational-age newborns. Female were 246 (46.5%) and male 283 (53.5%). The clinic attendance at the facility-based well-child clinic showed a decline from 98.1% at 6 weeks to 79.6% at 9 months of age. In parallel, pre-visit illness increased from 9.2% to 38.1%, infrequent feeding increased from 11.7% to 41.1%, and exclusive breastfeeding rate declined from 96.4% to 88.5%. Although 17% of the mothers in the cohort delayed initiating breastfeeding at birth, 96% of all these 529 mothers continued to breastfeed as at 9 months after birth. Cow milk supplementation was observed in 6.6% of cases, and complementary feeding with cow milk was noted in 38% of cases. Other milk feeds offered included soya and goat milk. The consumption of maize/millet/cassava porridge remained stable. Complementary family foods consisted mainly of carbohydrates (98.7%) and legumes (95.7%), as many families could not afford eggs or flesh foods. The weight z-score increments-over-time (velocity) significantly demonstrated earlier (4 weeks earlier) and higher weight faltering rates (22.6%) than the static (8.4-9.2%) or serial weight z-score methods of growth assessments. Infant weight deceleration was steepest during the age intervals between 6weeks and 14weeks. Weight z-score velocity plateaued between 6 and 9months of age. The mean weight increment percentages over the period of 9months for the small-for-gestational-age-born infants was 253% (sd 79), the appropriate-for-gestational-age-born infants was 172% (sd 48) and that for the large-for-gestational-age-born infants was 140% (sd 71), ANOVA p<0.001. However, when static measures were used to assess weight growth amongst these 3 categories of infants at 9months of age, the small-for-gestational age-born infants appeared to have the highest rate of underweight at 27.3%, while 7.6% of the appropriate-for-gestational-age-born infants were underweight. None of the large-for-gestational-age born infants was malnourished. The infant characteristics that significantly predicted postnatal weight deceleration were being born large-for-gestational age (OR=4.61[2.01, 10.59]) or male (OR=2.79 [OR=1.68, 4.62]). The small-for-gestational-age-born infants were 9.09times (95% CI 2.86, 33.33) more likely to experience weight acceleration, postnatally, compared to the other categories of infants.

**Conclusion:** This study highlights the considerable benefits (avoidance of mislabeling or failing to detect weight faltering) of utilizing weight increments or weight z-score velocity charts instead of static/serial measures for monitoring infant growth. It is essential to focus on the age intervals between 6 and 14 weeks after birth, male infants, large-for-gestational-age-born infants, previously ill infants, infant growth trajectories, types of feeds, and frequency of feeding during well-child clinic visits. Discouraging infant cow milk feeding practices is of utmost importance. Strengthening primary healthcare systems to enhance service delivery and increase contacts through home visits is imperative.

## Background

Promoting infant health within the critical “thousand-day window,” spanning from conception to a child’s second birthday, is paramount for ensuring survival.^1^ Globally, there has been a significant reduction in child mortality,^2^ dropping from 91 deaths per 1000 live births in 1990 to 43 in 2015.^3^ Notably, over a third of these deaths were attributed to undernutrition, particularly prevalent in developing nations. Providing preventive and promotive health care, through well-infant clinics, that involves regular monitoring of growth and development, screening for infectious diseases, administering essential vaccines, have contributed to this drop in infant mortality.^4^ Growth monitoring as an intervention, promotes early child development and is associated with long term health, economic, and social benefits.^5,6^

The growth monitoring program is extensively applied in numerous low-resource settings at the primary healthcare level, serving as a nutritional surveillance initiative linked to health promotion.^7^ This intervention entails at its core, monthly weight measurements and charting weight-for-age z-scores, identification of growth faltering, and counselling of caregivers on age-appropriate infant and young child feeding. These serial chartings are subsequently utilized to provide guidance to caregivers. The program fosters awareness regarding child growth and care practices, with the objective of enhancing demand for additional health services. The long-term ambition is to establish this initiative as a central component within an integrated child health system, thereby empowering caregivers to implement actions that cultivate growth-enabling environments for children.^7^ Growth monitoring program (GMP) services are usually offered alongside other child health services, including vaccinations, vitamin A distribution, free insecticide-treated bed net distribution, birth registration, education on disease prevention, and family planning encouragement. These GMP services are mainly delivered at child welfare clinics by community health nurses. The clinics operate as facility-based, community-based, and outreach clinics.^8^

Growth increments are influenced by the timing of measurements since growth occurs in a series of sporadic small or large spurts, which differ in intensity based on factors such as age, sex, developmental stage, and season. In infants, growth during the first six months is exponential rather than linear, according to American Academy of Pediatrics.^9^ Desch (1993) conducted a longitudinal study comparing incremental charts with standard growth charts for 42 high-risk infants and found that incremental weight charts were more effective than standard charts in visually representing weight changes over time.^10^

Exclusive breastfeeding (EBF) faces difficulties in maintaining adherence, even with mothers being informed about its significance.^11^ A more extended period of exclusive breastfeeding is associated with a lasting decrease in diarrhea-related gut microbiota imbalances, in addition to other advantages.^12,13^

The World Health Organization (WHO) and UNICEF advocate for a minimum exclusive breastfeeding coverage rate of 90% for infants in impoverished countries during their first six months.^14^ It is alarming that, even with breastfeeding rates exceeding 95% in Africa, inappropriate feeding behaviors, such as giving water and other liquids to infants, remain prevalent. These behaviors can undermine efforts toward exclusive breastfeeding and negatively affect infant health and nutrition. Despite initiatives led by organizations like the WHO and UNICEF, along with local governments and NGOs, considerable obstacles continue to hinder the achievement and maintenance of high exclusive breastfeeding rates. Moreover, the elevated levels of bottle-feeding in certain nations (over 30% in Tunisia, Nigeria, Namibia, and Sudan) intensify the problem, as it may lead to reduced exclusive breastfeeding rates and potential health issues for both mothers and infants.^15,16,17,18^

Suboptimal utilization of the GMP services can lead to significant negative effects on different aspects of the society. Therefore, this research aimed to assess the clinic attendances, feeding patterns, including rates of exclusive breastfeeding, alternative feeding practices, and growth patterns in 529 infants visiting a well-child clinic in a rural community of East Africa.

## Materials and methods

The methods employed in carrying out this study is discussed in this section.

## Study setting

Rwanda is a low-income, agricultural and landlocked country with approximately 11 million people living in five provinces, covering an area of 26,338 km.^19^ It is called the home of a ‘1000 hills’.^19^ The limited area of flat land available in most part of Rwanda is a hindrance to farming, animal rearing and construction of standard residential houses, among others.

Rwanda has an average of 4.4 persons per household and a gross domestic product per capita of US $780.80. About half (48%) of its population were under 19 years of age and 39% lived below the poverty line with a life expectancy at birth of 71.1 years for women. Literacy rate among 15– 49 years old women was at 80%. In addition, 87.3% of the population have health insurance and access to health services; spending an average of 47.4 min to reach a health centre.

Gitwe village is located on a high altitude of 1,674 meters above sea level, in the southern province, 240km from Kigali, which is the capital city of Rwanda. Gitwe general hospital began in 1995, immediately after the genocide, for the purposes of providing medical services and later training to this isolated community. The hospital currently has 100% government support, since year 2020. The maximum number of deliveries at the hospital per month was about 200 and about 50 infants visit the well-child clinic per day. There are only 2 immunization days per week.

Some of the challenges in the hospital include poor specialist coverage, few trained health workers, poor supply of equipment, water, electricity, laboratory services and medicines. Accessibility to the health facility along the hilly, sometimes unmotorable untarmacked roads can be extremely challenging, especially at the 2 peaks of the rainy seasons, in June and October.

Challenging cases are referred to the University of Rwanda Teaching Hospital in Butare or Kigali. Gitwe village was selected for this study, because there was no published birth or infant data from this poorly researched, remote, relatively inaccessible community. In 2019, birth and up to 9 months of age, feeding and growth data on 529 mother-singleton newborn pairs were compiled in this village, over a period of 12 months in the delivery, postnatal wards and clinics of Gitwe general hospital and at its well-child clinic.^19^

## Data source and sample

This was a secondary analysis of data collected for a prospective cohort study on growth faltering over a period of 12 months, until infant was 9 months old. Sample size of 529 was calculated using the Epitool sample size calculator.^20^ Mother-newborn pairs were recruited consecutively, on first-come-first-serve basis at birth. Newborn gestational age was determined using the record of the maternal first date of the last menstrual period (LMP), fetal ultrasound dating and/or expanded new Ballard criteria. Newborn and infant weight (kg) were measured and recorded to the nearest decimals, from birth. The birth/weight-for-age z-scores (WAZ) and percentiles (WAP) were computed using the PediTool calculator.^21,22^ Normal intrauterine growth was defined as birth weight-for-gestational-age percentile between 10^th^ and 90^th^ percentile; and with this definition, newborns were classified into small-for-gestational-age, appropriate-for-gestational-age and large-for-gestational-age. Weight deceleration/catch down, post-delivery was considered, if the birthweight-gestational-age z-score had fallen by more than 0.67points over any 2 time intervals. NICE and WHO weight-for-age percentile of ≤2 and z-score <-2 at any single visit, was considered as static weight faltering, respectively.^22^ Infant weight percent increments were also calculated. Mothers were interviewed using standardized infant feeding questionnaires^23^ and the food types were classified into 8 groups: (A) Breastmilk; (B) Grains, roots, tubers and plantains; (C) Pulses (beans, peas, lentils), nuts and seeds (D) Dairy products (milk, infant formula, yoghurt, cheese); (E) Flesh food (meat, fish, poultry, and organ meats); (F) Eggs; (G) Vitamin A-rich fruits and vegetables; (H) Other fruits and vegetables^24^ at every clinic visit. The questionnaires were read to the mothers and filled by the research assistants.

To ensure the quality of data collected, 2 registered nurses were trained as research assistants at Gitwe Hospital on the over-all procedure of newborn anthropometry and data collection by the investigator. The questionnaires were pre-tested before the main data collection phase, on 10 mother-infant pair participants (2% of the total sample). The investigator closely followed the day-to-day data collection process, to ensure completeness and consistency of the questionnaires administered each day, before data entry.

## Statistical analysis

Data clean up, cross-checking and coding were done before analysis. These data were entered into Microsoft Excel statistical software for storage and then exported to Statisty-free web-based online statistics app^25^ for further analysis. Both descriptive and analytical statistical procedures were utilized. Participants’ categorical characteristics were summarised in frequencies, percentages figures. Chi test was used to determine association between infant characteristics and weight faltering. The outcome variables (weight (kg), weight z-scores, weight z-score differences and weight percent increase) were essentially normal in their distribution and hence, parametric tests such as t-test, ANOVA were performed to determine relationship between infant characteristics and infant weight parameters. Logistic regression analysis was performed to determine the infant factors that predict weight deceleration and generate Odds ratio with 95% confidence interval. For all, statistical significance was declared at p-value <0.05. The reporting in this study were guided by the STROBE guidelines for observational studies.^26^

## Ethics

The Health Sciences Research Ethics Committee of the University of the Free State in South Africa gave the ethical approval for the collection of the primary data for the original study-’Growth and growth faltering in a birth cohort in rural Rwanda: a longitudinal study’ with Ethical Clearance Number: UFS-HSD2018/1493/2901. Written permission to collect data was obtained from the Director of Gitwe Hospital, Rwanda. Verbal consents were obtained from eligible mothers as they were recruited consecutively from the postnatal ward after delivery, with the attending nurse as a witness. The content of the research information booklet and consent letter were read out to the mothers and their permission was sought after ensuring that they understood the purposes, methods, pros and cons of the research. The participants were given research identity numbers and the principal investigator was responsible for the safe keeping of the completed questionnaires and collected data, to ensure anonymity and confidentiality of the participants.

## Results

The following are the results obtained from the study.

## Participants

Five hundred and ninety-seven (597) newborns were delivered at Gitwe Hospital, Rwanda, between 22^nd^ January and 9^th^ May 2019, out of which, eligible 529 mother-newborn pairs were enrolled into the prospective cohort study from birth, Figure 1. At birth, there were 113 (21.4%) small-for-gestational-age, 379 (71.6%) appropriate-for-gestational-age and 37(7.0%) large-for-gestational-age newborns. Female were 246 (46.5%) and male 283 (53.5%).^27^ Growth and feeding data were collected from these 529 infants from birth to the age of 9months at the maternal and well-child clinic.

**Fig 1:**
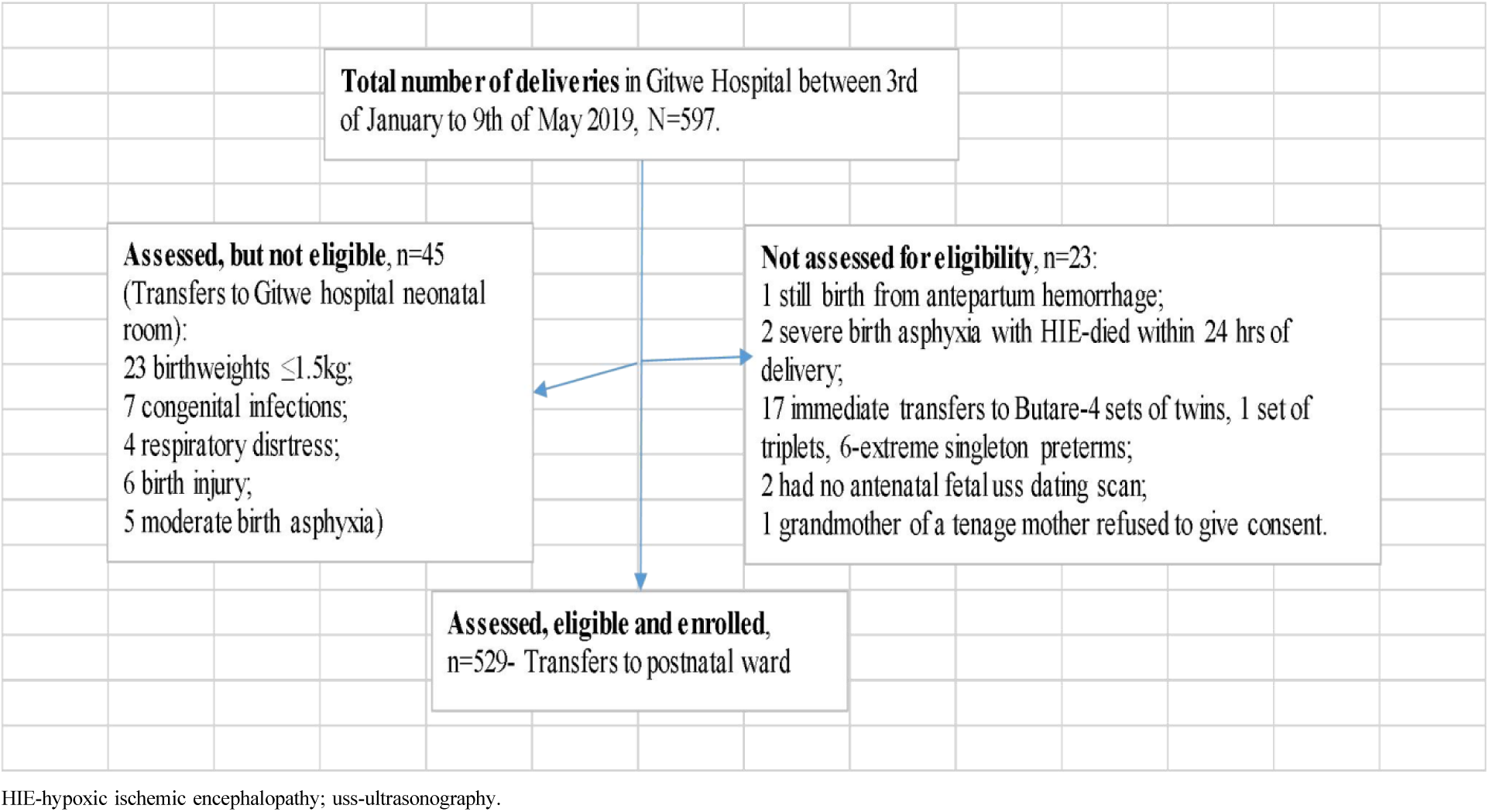
Flow of participants from admission to recruitment into study.

## Pre-visit illness and attendance at the Maternal and Child Health Clinic

Attendance at the clinic declined as observed from the increasing rates of absenteeism (10 [1.9%] at 6 weeks, 62 [11.7%] at 10 weeks, 15 [2.8%] at 14 weeks, 40 [7.6%] at 6 months and 108 [20.4%] at 9 months). As such only 292 (55.2%; 151[51.7% female] 141 [48.3% male]) of the 529 newborns recruited from birth had regular and complete attendances at all the 5 clinic visits, Table 1. Majority (98.3%) of these regular attendees were appropriate-for-gestational age babies. Infants that had pre-visit illness were 47 (9.2%), 54 (11.6%), 77 (15.9%), 75 (15.3%) and 149 (38.1%) at 6weeks, 10weeks, 14weeks, 6months and 9months of age respectively.^28, 29^ The 2 most common illnesses were respiratory and diarrhea diseases.

**Table 1:** Follow-up visits, n =529.

| Number of follow up visits | Frequency | Percentage |
| --- | --- | --- |
| 1 (6 week) | 4 | 0.8 |
| 2 (10 week) | 8 | 1.5 |
| <sup>31</sup>3 (14 week) | 39 | 7.4 |
| 4 (6 month) | 186 | 35.2 |
| 5 (9 month) | 292 | 55.2 |

## Feeding

Of the 529 newborns recruited at birth, 83% were fed within 1 hour (37.6% of the babies delivered by cesarean section and 4% of those delivered vaginally received delayed feeds). Prelacteal feeds were offered to 6% of these 529 babies at birth (12.7% of babies delivered by cesarean section and 1.9% of babies delivered vaginally). ^27^

Exclusive breastfeeding rate declined from 96.4% at 6 weeks, 96.3% at 10 weeks, 83.9% at 14weeks^30^ to 88.5% at 6months of age. Continued breastfeeding was observed in 96% of the infants at 9months of age.

Infants receiving infrequent feeding (fed <8 times a day) were 57 (11.7%) of the 489 infants that visited the clinic at 6 months. Infrequent feeding increased to 41.1% at 9 months. Supplementary and complementary liquid feeds offered to infants in the cohort are shown in Table 2, with home modified cow milk being the most common, followed by soya milk. Goat milk was offered after 6 months.

**Table 2:** Frequency and Types of alternate liquid feeds (n=529)

|  | Type of alternate liquid feeds | Up to 6 months of age (%) | After 6 months (%) |
| --- | --- | --- | --- |
| 1 | Home modified cow milk | 35 (6.6) | 201 (38.0) |
| 2 | Soya | 13 (2.5) | 107 (20.2) |
| 3 | Porridge (millet/maize/cassava) | 16 (3.0) | 54 (10.2) |
| 4 | Formula | 2 (0.4) | 3 (0.6) |
| 5 | Goat milk | ---- | 2 (0.4) |

Complementary feeds offered to the infants were in semi-solid or marshed, soft, solid forms, Table 3. Most common was food group B (grains) in form of liquid or stiff porridge.

**Table 3:**
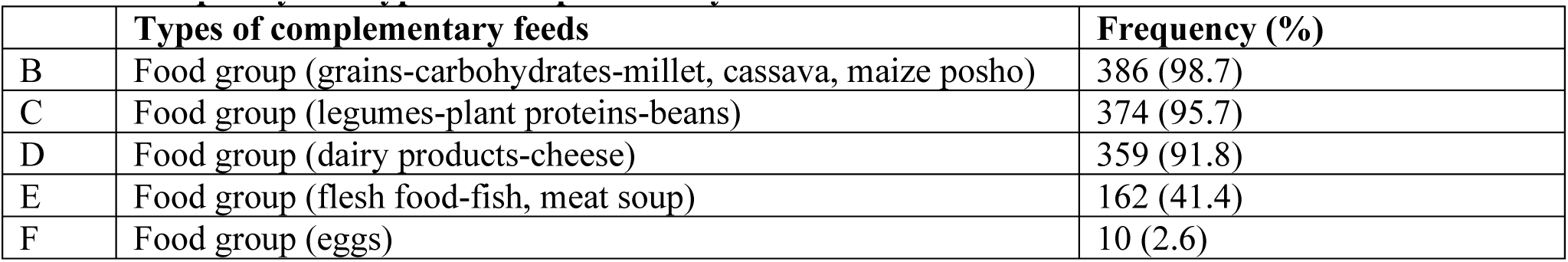
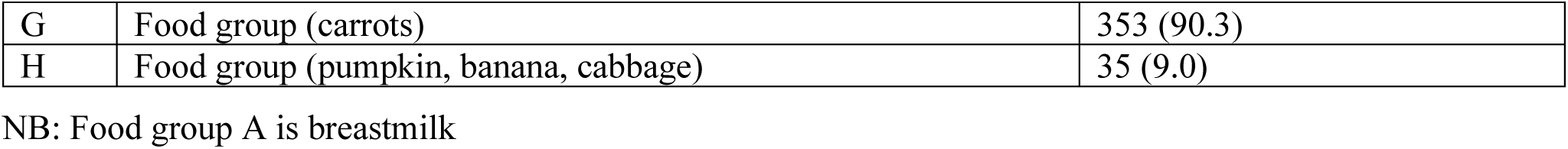
Frequency and types of complementary feeds.

|  | Types of complementary feeds | Frequency (%) |
| --- | --- | --- |
| B | Food group (grains-carbohydrates-millet, cassava, maize posho) | 386 (98.7) |
| C | Food group (legumes-plant proteins-beans) | 374 (95.7) |
| D | Food group (dairy products-cheese) | 359 (91.8) |
| E | Food group (flesh food-fish, meat soup) | 162 (41.4) |
| F | Food group (eggs) | 10 (2.6) |
| G | Food group (carrots) | 353 (90.3) |
| H | Food group (pumpkin, banana, cabbage) | 35 (9.0) |

NB: Food group A is breastmilk

## Weight assessment of the infant cohort

The weight of the infant cohort were assessed at birth, 6 weeks, 10 weeks, 14 weeks, 6 months and 9 months of age. The cohort doubled its birthweight at 10weeks, and almost tripled it by 9months of age, Table 4.

**Table 4:** Weight (kg) measurements and z-scores of infants from birth to 9 months of age.

| COHORT |  |  |  |  |  |  |  |  |
| --- | --- | --- | --- | --- | --- | --- | --- | --- |
| Age |  | Weight (kg) |  | Weight z-score |  | Weight z-score increments |  |  |
|  | n | Median | IQR | Median | IQR | n | Median | IQR |
| **Birth** | 529 | 3.1 | 2.8, 3.5 | -0.5 | -1.2, 0.3 |  |  |  |
| **6 weeks** | 512 | 5.5 | 4.9, 6.0 | 0.8 | -0.0, 1.5 | 512 | 1.3 | 0.4, 2.0 |
| **10 weeks** | 466 | 6.5 | 6.0, 7.0 | 0.9 | 0.2, 1.6 | 457 | 0.0 | -0.7, 0.9 |
| **14 weeks** | 483 | 7.1 | 6.3, 7.9 | 0.8 | -0.2, 1.6 | 432 | -0.0 | 1.1, 0.8 |
| **6 months** | 489 | 8.0 | 7.2, 8.8 | 0.4 | -0.5, 1.2 | 454 | -0.5 | -1.4, 0.9 |
| **9 months** | 391 | 8.5 | 7.9, 9.3 | -0.0 | -0.9, 0.8 | 359 | -0.4 | -1.6, 0.6 |

**Table 2:** Gender-specific weight of infants from birth to 9 months of age.

|  | FEMALE |  |  |  |  | MALE |  |  |  |  |
| --- | --- | --- | --- | --- | --- | --- | --- | --- | --- | --- |
| Age | Weight (kg) | | | Median weight z (IQR) | Median weight z $\hat{\Delta}$ (IQR) | Median weight z$\hat{\Delta}$ (IQR) | Median weight z (IQR) | Weight (kg) | | |
|  | n | Median | IQR |  |  |  |  | n | Median | IQR |
| Birth | 246 | 3.0 | 2.8, 3.4 | -0.5 (-1.2, 0.2) |  |  | -0.5 (-1.2, 0.3) | 283 | 3.2 | 2.9, 3.5 |
| 6wks | 242 | 5.5 | 5.0, 6.0 | 1.0 (0.3, 1.6) | 1.6 (0.7, 2.3) | 1.0 (0.2, 1.7) | 0.6 (-0.3, 1.4) | 270 | 5.5 | 4.9, 6.0 |
| 10wks | 219 | 6.4 | 5.9, 6.9 | 1.1 (0.3, 1.7) | -0.0 (-0.8, 0.8) | 0.1 (-0.6, 1.1) | 0.8 (-0.1, 1.3) | 247 | 6.5 | 6.0, 7.0 |
| 14wks | 234 | 7.0 | 6.0, 7.7 | 1.0 (0.1, 1.8) | -0.1 (-1.2, 0.6) | 0.0 (-0.9, 1.0) | 0.7 (-0.5, 1.4) | 249 | 7.4 | 6.4, 8.0 |
| 6mths | 236 | 8.0 | 7.3, 8.9 | 0.7 (-0.0, 1.5) | -0.2 (-1.1, 1.1) | -0.7 (-1.5, 0.6) | -0.0 (-0.9, 0.9) | 253 | 8.0 | 7.2, 8.7 |
| 9mths | 188 | 8.5 | 7.7, 9.4 | 0.3 (-0.6, 1.0) | -0.6 (-1.7, 0.4) | -0.3 (-1.4, 0.9) | -0.4 (-1.1, 0.4) | 203 | 8.5 | 8.0, 9.3 |

The median weight of the cohort, progressively increased all through the study, but there was a progressive drop in the median weight z-scores from the 10^th^ week to 9 months of age. However, the median increment weight z-scores curve began its persistent decline, earlier and much lower than the other weight curves, from 6 weeks of age, Fig. 2. The difference in weight z-scores was positive and highest between birth and 6 weeks of age; lowest between 6 and 10weeks of life; and plateaued between 6 and 9months of age. Weight faltering was at 5.7% (30) at birth, 2.9% (15) at 6 weeks, 2.4% (11) at 10weeks, 5.6% (27) at 14weeks, 2.7% (13) at 6 months and 8.4% (33) at 9 months of age, using static weight-for-age percentile <2 as cut-off point. However, when weight z score-increment, from birth to 9months of age was assessed among the 292 infants that completed 5 clinic visits, the percentage of those that were faltering was 22.6% (66 out of 292). When cut-off point of –2 z-score was applied, weight faltering rate was 9.2% (27 out of 292) at 9 months of age. Using chi^2^ test, there was a significant difference (Chi value 33.02, p<0.001) in the percentage of weight faltering obtained from the 2 different methods (static versus incremental) of weight faltering assessment.

**Fig. 2:**
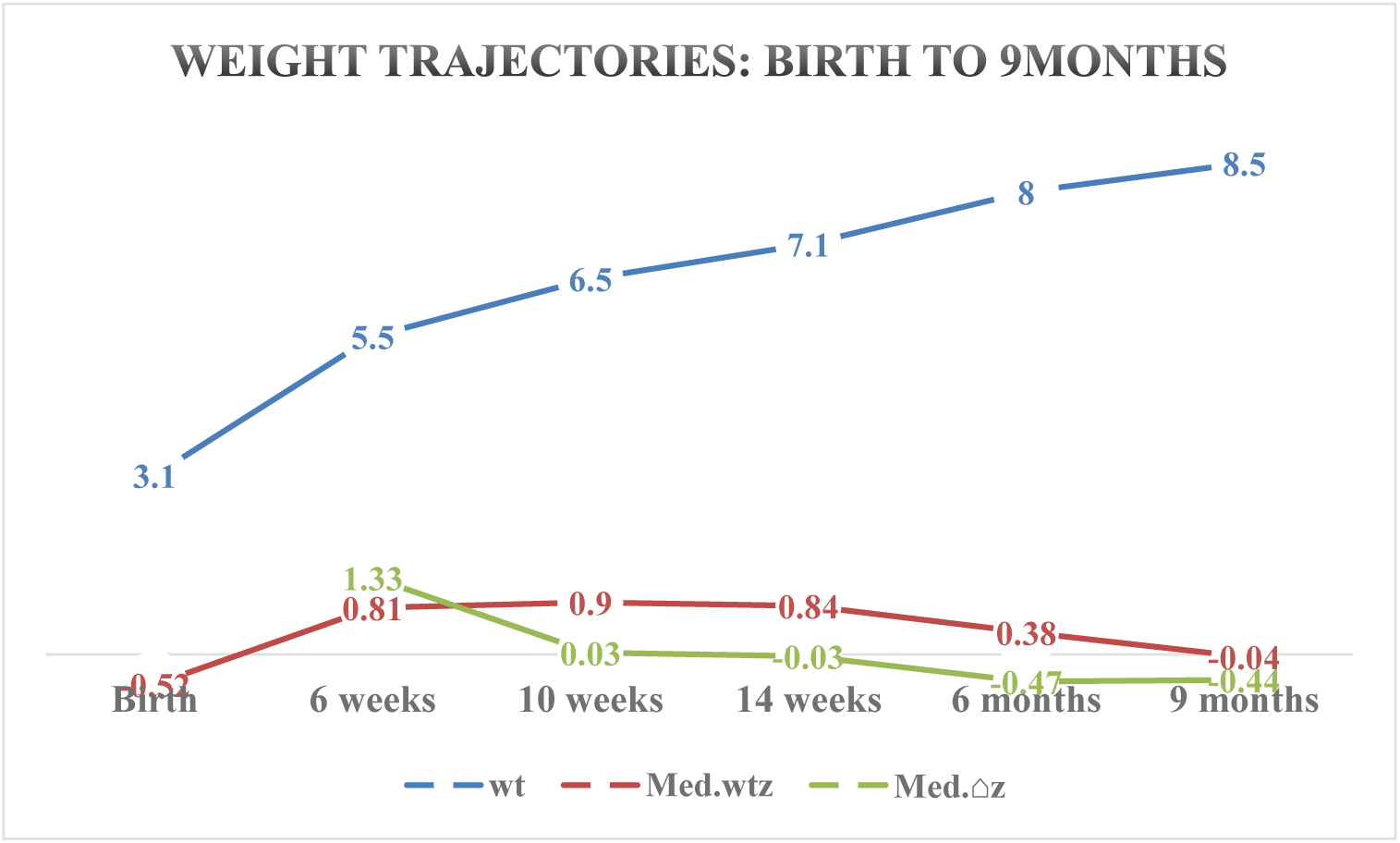
Trajectories of median-weight, weight z-score and weight z-score increments from birth to 9months of age.

## Infant weight and sex

The median weight, z-scores and z-score increments over the period from birth to 9months of age for the girls and boys in the cohort is shown in Table 2. The boys were bigger in size than the girls at most visits except at 6 months of age. There was a significant difference in the weight z-scores increments between the male and female infants, p=0.001. Weight z-score increment in the age intervals were significantly higher in the female than male infants (mean difference Male-Female was –0.1, SE 0.03, t –3.22, p=0.001), even though no difference was found when the male-female weight (kg) were compared (mean difference Male-Female was 0, SE 0.08, t 0.02, p=0.986). Weight percentage increment analysis from birth to 9months of age revealed a mean weight increase of 172% (sd 57) for boys and 179% (sd 52) for girls, p=0.209, Figure 3.

**Figure 3:**
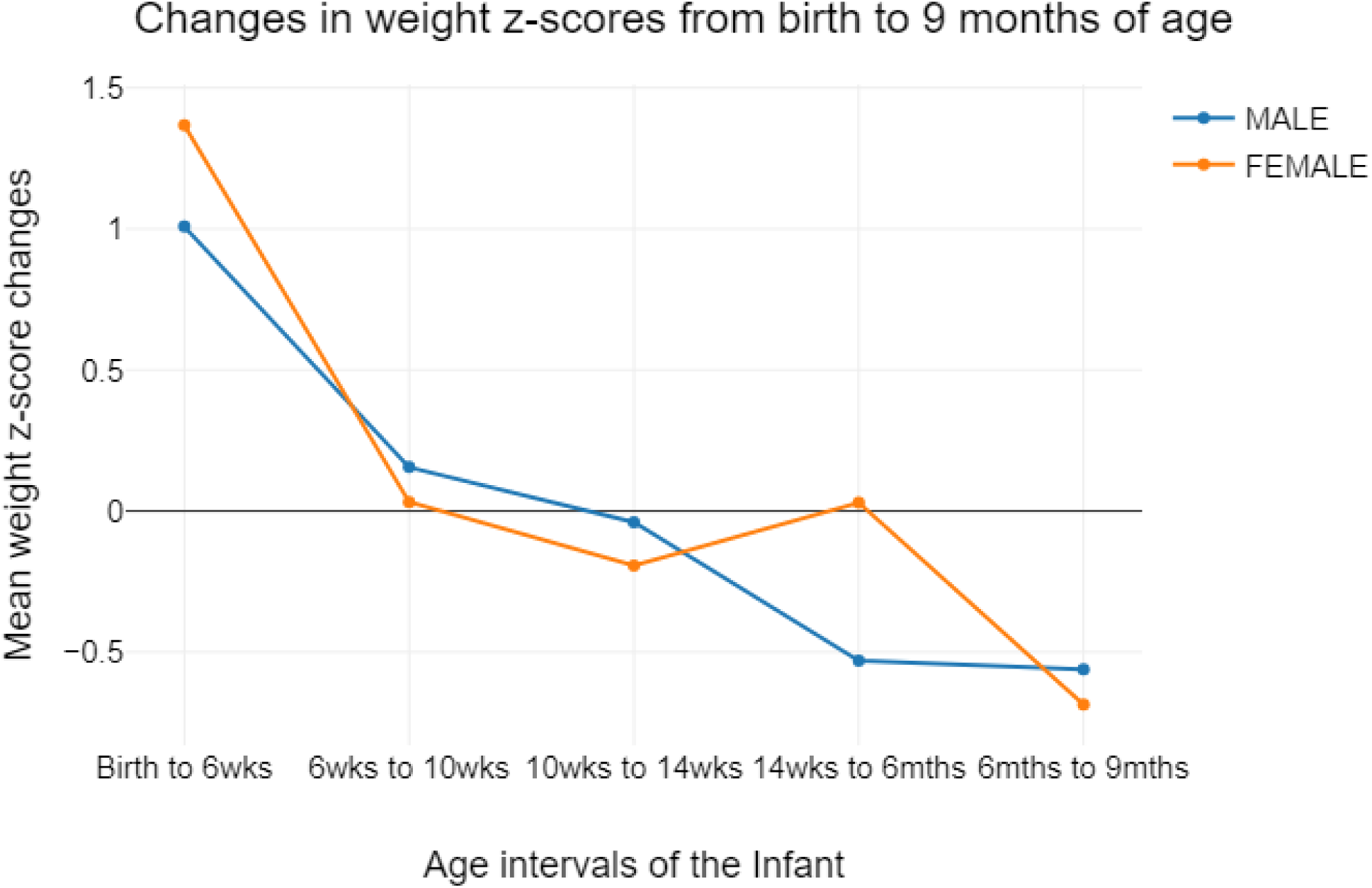
Mean weight z-score changes by sex from birth to 9months of age.

A two-factor analysis of variance with repeated measures was performed for the infants (292 in number) with complete attendance at all the clinic visits and showed that there was a significant difference in the weight z-score increments between age intervals of “Birth to 6wks, 6wks to 10wks, 10wks to 14wks, 14wks to 6mths and 6mths to 9mths” in the cohort, p=<0.001. The weight z-score increment in the interval from birth to 6weeks of age was significantly higher; while that from 6months to 9 months of age was significantly slower than any other age interval, according to Bonferroni Post-hoc test.

## Infant intrauterine growth and postnatal weight gain

The mean weight increment percentages over the period of 9months for the small-for-gestational-age-born infants was 253% (sd 79), the appropriate-for-gestational-age-born infants was 172% (sd 48) and that for the large-for-gestational-age-born infants was 140% (sd 71), ANOVA p<0.001. However, when using static measures to assess weight growth amongst these 3 categories of infants at 9months of age, the small-for-gestational age-born infants appeared to have the highest rate of malnutrition at 27.3%, while 7.6% of the appropriate-for-gestational-age-born infants were malnourished. None of the large-for-gestational-age born infants was malnourished.

## Logistic regression analysis of infant factors that predict weight deceleration

Logistic regression analysis was performed to examine the influence of being born large-for-gestational-age, small-for-gestational-age or male on postnatal weight deceleration. Logistic regression analysis shows that the model as a whole was significant (Chi2(3) = 62.35, p <.001, n = 391).

The coefficient (b) of the variable ‘large-for-gestational-age’ was 1.53, which was positive. The odds ratio was 4.61. This means that, if the infant was born large-for-gestational-age, the probability of weight deceleration increases 4.61 times. The p-value was <0.001 and this indicates that this influence was statistically significant.

The coefficient (b) of the variable ‘small-for-gestational-age’ was –2.25, which was negative. The odds ratio was 0.11. This means that if the infant was born small-for-gestational-age, the probability of weight deceleration decreases 9.09 times. The p-value of <0.001 indicates that this influence was statistically significant.

The coefficient (b) of the variable ‘male’ was 1.03, which was positive. The odds ratio was 2.79. This means that if the infant was born male, the probability of weight deceleration increases 2.79 times, compared to their female counterpart. The p-value of <0.001 indicates that this influence was statistically significant.

## Discussion

This study examined the clinic attendances, feeding practices and weight growth patterns of 529 infants recruited from birth to 9months of age at the well-child clinic in a rural community.

Clinic attendance rates at both the 10-week and 9-month clinic visits were noted to be at their lowest levels, with only 55.2% of this cohort maintaining consistent attendance. ^27,28,29,30^ This regularly attending group primarily comprised infants born appropriate-for-their-gestational-age (AGA). In contrast, the small-for-gestational-age (SGA)-born infants—who are expected to encounter greater health challenges—exhibited irregular attendance patterns. This is particularly troubling as it leads to missed opportunities for early identification and intervention for childhood illnesses, as well as for ensuring that children receive necessary immunizations and counseling. The rising incidence of illness as these infants age, underscores the urgency of regular healthcare access, as SGA infants may experience severe health outcomes, potentially leading to fatalities at home without the knowledge of healthcare providers. This irregular clinic attendance could be attributed to several factors, including the difficulties posed by unpaved, hilly roads, particularly during peak rainy seasons, when travel becomes significantly more complicated. Additionally, scheduling conflicts arising from market days, farming activities, and clinic visits can adversely affect attendance. Many women are the primary earners in their households, making it challenging for them to prioritize clinic visits.^31^ Consequently, mothers who are constrained by work or market obligations may find it increasingly difficult to endure the delays associated with long queues of patients at the clinic.

Missing clinic attendances represent a significant global concern, even in developed nations such as the United States, where children frequently miss between one-third to one-half of their recommended well-child appointments.^32^ Increased attendance at well-child clinics has been associated with enhanced infant growth and a reduction in the utilization of emergency services and hospitalizations. ^33, 34,35^

The current study corroborates these findings, revealing that as clinic visits, exclusive breastfeeding rates, and frequency of feedings decline, the incidence of infant illness and weight faltering rises. Among those who consistently attended the clinics, a greater number of female infants were observed, and these girls demonstrated superior growth compared to their male counterparts in this Rwanda study.

Several factors have been linked to suboptimal attendance at well-baby clinics in other studies, including socioeconomic status, race, young maternal age, single marital status, higher parity, lower educational attainment, mental health challenges, transportation issues, and inadequate insurance coverage. Furthermore, mothers who received sufficient antenatal care exhibit higher adherence to well-child clinic visits, as similar factors influence both types of medical appointments. ^33,34^

To improve attendance rates at post-delivery clinics, it is imperative to implement strategies such as enhancing road infrastructure, conducting public education campaigns, offering flexible clinic scheduling, and providing home visits. It is critical to address these matters, as neglecting medical issues during this essential period of postnatal life can have significant implications for both mothers and infants.

Despite the challenges associated with rural living, 96% of the infants in this cohort continued to breastfeed by the age of 9 months. The rate of exclusive breastfeeding (EBF) experienced a decline from 96.4% at 6 weeks,^28^ 96.3% at 10 weeks,^29^ and 83.9% at 14 weeks,^30^ eventually reaching 88.5% at 6 months of age. Although the EBF rates are decreasing, they remain higher than the global targets of 50% by 2025 and 70% by 2030.^36^ It is worth noting that breastfeeding prevalence is notably greater in low-income and middle-income countries compared to high-income countries. Furthermore, these EBF rates in this current study, significantly surpass the 59.2% average reported for infants aged 0-5 months in the East African region, as outlined in the 2022 Global Nutrition Report.^37^

In this cohort, the supplementary milk or liquid feeds administered to infants prior to 6 months of age, primarily included modified cow milk, soy milk, and maize/cassava/millet porridge. This practice can be attributed to the fact that nearly every family in the village maintains a backyard garden, allowing for the cultivation of staple foods and the keeping of livestock. The availability of infant formula was significantly limited within this economically disadvantaged community, with consumption rates documented between 0.4% and 0.6% in this study. This study observation aligns with the findings of Neves et al. (2021).^36^ Neves et al. reported an increase in formula consumption during the first 6 months of life in upper-middle-income countries, as well as in regions such as East Asia, the Pacific, Latin America, the Caribbean, the Middle East, North Africa, Eastern Europe, and Central Asia. In contrast, the rates of infant formula consumption in sub-Saharan Africa and South Asia have remained below 8%.

Although Neves et al. reported a global decline in animal milk consumption, the current study indicates an increase in cow milk intake, rising from 6.6% at 6 months to 38% at 9 months.^36^ This finding aligns with a similar cross-sectional study conducted in 2021 across five health facilities in Kigali, the capital of Rwanda, which involved 221 mothers and their infants. In that study, 47.1% of mothers supplied infant formula, 20.4% provided cow’s milk, and 14.5% offered porridge, among other supplementary feeds. The rates of exclusive breastfeeding (EBF) were substantially lower in the Kigali study,^38^ with EBF rates of 85.1% at birth, 81.9% at 3 months, and 57.5% at 5 months, compared to this rural study, which reported rates of 94% at birth, 96.4% at 6 weeks, 96.3% at 10 weeks, and 88.5% at 6 months. These discrepancies suggest a higher socio-economic status among mothers in the Kigali study than those in the rural areas. Notably, mothers in the Kigali study began introducing supplementary feeds as early as 1 month of the infant’s age.^38,39,40^

The findings from both studies conducted in Rwanda illustrate the prevalent practice of feeding infants with cow milk in both urban and rural settings. It is essential to recognize that economic constraints in rural areas often lead mothers to provide unmodified cow milk to their infants. Research has shown the detrimental effects of cow milk on infant health, including the potential development of iron deficiency anemia due to gastrointestinal bleeding, the inhibition of non-heme iron absorption owing to its high calcium and casein content, and its insufficient iron content. Iron is a critical nutrient for optimal neurodevelopment within the first year of life.^41,42^

The consumption of soya milk in this study increased notably as both a supplementary and complementary dietary option, rising from 2.5% to 20.2%. This level of soya consumption parallels the 20% observed among infants in Canada in 2009. However, the soya milk utilized is often prepared at home and is vulnerable to contamination due to inadequate water supply in the community, contributing to the incidence of diarrhea reported in this study.

It is essential to recognize that soya milk is not a suitable choice for preterm infants or those born SGA, primarily due to its elevated levels of phytoestrogens, aluminum, phytates, and calcium. These compounds can disrupt thyroid function and impede the absorption of iron, calcium, and other essential nutrients, potentially resulting in anemia and inadequate bone development in these infants, as noted by the Canadian Paediatric Society. Soya milk should only be recommended for infants who are unable to consume breast milk or dairy-based options for health, cultural, or religious reasons, such as adherence to a vegan lifestyle or the presence of galactosemia.^43,44^

Furthermore, a small percentage of infants in this cohort received goat milk as a complementary feed, which is known to be deficient in folate. Additionally, these infants typically do not receive any vitamin or iron supplementation from health clinics, which may further aggravate nutritional deficiencies.^41,42^

After 6 months, these infants were transitioned to a diet primarily consisting of carbohydrates and legumes, which are more economically accessible than eggs, meat, and fish. Despite this inadequate nutritional quality, this cohort demonstrated substantial weight increase, doubling its birth weight by 10 weeks and tripling it by 9 months of age. Nonetheless, the rate of weight faltering exhibited variability, peaking at 9 months.

A diet rich in carbohydrates typically offers lower nutritional value compared to breast milk and is significantly associated with an increased risk of overweight in infants. Poverty was identified as a major factor contributing to the decline in breastfeeding practices and poor complementary feeding habits, as illustrated in a qualitative study conducted by Ahishakiye et al. (2019) in Muhanga, a rural town in Rwanda in the same province with the current study site. As a result, despite possessing adequate knowledge of nutrition, some parents chose to sell more nutritious foods, such as eggs, in order to purchase less expensive alternatives like maize.^31^

The findings from the current study suggest that the use of weight z-score increments-over-time intervals (velocity) provides more insightful data than static assessments, as it identified instances of early weight faltering that were not detected by other methods. Furthermore, static assessments were ineffective in distinguishing differences in infant growth when compared to incremental or interval growth evaluations.

According to Panpanichi et al.,^43^ existing methods for growth assessment—relying on static weight z-scores or longitudinal serial weight z-scores that monitor deviations from the mean—may prove inadequate for identifying genuine weight faltering promptly. This limitation arises because many infants transition across percentile channels (both upward and downward) on growth charts, which complicates healthcare providers’ ability to accurately identify growth issues. In contrast, the analysis of weight z-score differences over time intervals or the application of weight velocity standards demonstrated distinct advantages, as evidenced by the significant variations in weight patterns observed in this study. Specifically, weight velocity (measured through weight z-score differences) revealed faltering 4 weeks earlier and with a greater magnitude (22.6% vs. 9.6%) compared to static and serial weight z-scores in this current study. This aligns with the 24% period prevalence rate of weight faltering reported by Ross et al. in 2009,^44^ derived from a cohort of 3,727 infants in Colorado, USA, over a duration of 6 months, using weight velocity method.

Additionally, insights from this study indicate a propensity to misclassify small-for-gestational-age (SGA) infants as faltering while failing to identify faltering in large-for-gestational-age (LGA) infants when solely relying on static growth measures. Such misclassification may lead to excessive interventions for SGA infants, potentially resulting in cardiovascular and metabolic disorders later in life.^44^ The inability to recognize weight faltering in LGA infants at an early stage, due to their seemingly ‘large’ appearance, may lead to a considerable missed golden opportunities for timely intervention. Such oversight could facilitate the progression of underlying pathology, ultimately resulting in the development of overt malnutrition along with its associated short-term and long-term sequelae.

The analysis of weight growth patterns, as indicated by weight z-score differences over specified time intervals, was informed by a systematic review and meta-analysis of studies conducted across 30 countries in 2020. Findings indicated that boys exhibited a greater susceptibility to undernutrition, particularly manifesting as wasting, underweight, and stunting, compared to girls, as evidenced by 59 out of the 74 studies examined. While this study also found that girls were more likely to experience wasting, this observation lacked statistical significance. Furthermore, the authors noted that these sex differences in undernutrition were most pronounced during the first 30 months of life, but tended to diminish as children aged. This phenomenon may be attributed to both biological factors, including immune and hormonal influences, as well as social determinants.^45^

Despite an observed increase in the rate of weight faltering among infants in this cohort, the figures remained significantly lower than the 50.4% reported for 4-month-old infants in an urban Sri Lankan setting, where the exclusive breastfeeding rate at 4 months was 88%.^41^ This discrepancy underscores the urgent need for interventions to address the declining rates of exclusive breastfeeding within this rural, socio-economically disadvantaged community.

Therefore, it is imperative to closely monitor male infants, those who have been previously ill, along with their growth trajectories, type of feeding, and frequency of feeding during each clinic visit.

## Limitations

One of the limitations of this study was the loss to follow-up of infants at the child health clinic. Additionally, the exclusion of small and sick babies from the study at birth may have artificially lowered the infant weight faltering rate in the cohort. The authors acknowledge the various definitions of infant weight faltering used in other studies, highlighting the absence of global consensus on standard weight faltering definitions for infants below the age of 6 months.

## Strength

As a result of the longitudinal nature of the parent research, this article presented age-specific weight growth statistics for infants from birth to 9 months of age, which are difficult to obtain in current literature.

## Conclusion and Recommendations

This study is emphasizing the greater advantages of using weight z-score velocity charts rather than static measures to monitor infant growth. Close attention must be paid to the age intervals between 6 and 14 weeks after birth, male infants, previously ill infants, infant growth trajectories, types of feeds and frequency of feeding during the well-child clinic visits. Infant cow milk feeding practices must be strongly discouraged. There is need to strengthen the primary healthcare systems to improve service delivery and increase contacts through home visits.

## Data Availability

All data produced in the present study are available upon reasonable request to the authors

## Author Contributions

The corresponding author (Dr Adenike Oluwakemi Ogah) conceived and designed the study, collected data and conducted data analysis, interpreted the results, and drafted the manuscript.

## Acknowledgements

The author is extremely grateful to the participants involved in this study, to the staff of Gitwe Hospital and clinic in Rwanda and to the research team.

